# Rivaroxaban is Comparable to Warfarin in Prevention of Thromboembolism in Patients with Non-Valvular Atrial Fibrillation with Valvular Heart Disease: A Systematic Review and Meta-analysis

**DOI:** 10.1101/2021.07.08.21260233

**Authors:** Nischit Baral, Anjan Katel, Govinda Adhikari, Mahin R. Khan, Hafiz M. W. Khan, Rohit Rauniyar, Maxwell Akanbi, Bilal Malik, Muhammad Ahmad, Ashiya Khan, Basel Abdelazeem, Pramod Savarapu, Sakiru O. Isa, Arvind Kunadi, Parul Sud, Hameem U. Changezi

**Author notes:** Address for correspondence: Nischit Baral, MD, Internal Medicine Residency: McLaren Flint/Michigan State University College of Human Medicine, 401 S Ballenger Hwy, Flint, MI, 48532, OR. Funding: None. Disclosure: All authors have no conflict of interest to disclose.

## Abstract

**Objectives:** While the use of novel oral anticoagulants (NOACs) has been approved in the treatment of non-valvular atrial fibrillation (NVAF), we are lacking studies on individual NOACs in NVAF with valvular heart disease (VHD) including bio-prosthetic valve and valve repair. We aimed to determine the efficacy and safety of rivaroxaban compared to warfarin in prevention of thromboembolism in patients with NVAF with VHD.

**Methods:** We searched PubMed, MEDLINE, and EMBASE including only RCTs and Cohort studies from inception till April 2021. Eligible studies compared rivaroxaban with warfarin in patients with NVAF with VHD. We excluded patients with valvular AF. We used Review Manager (version 5.4, Cochrane Collaboration, Oxford, UK) applying the Mantel-Haenszel test and followed PRISMA guidelines. Risk ratio (RR) and 95% confidence intervals (CIs) were estimated using a random-effects method and heterogeneity using I squared test.

**Results:** We had total of 23136 participants in both groups. Our results showed stroke and systemic thromboembolism in 88 of 4258 (2.06%) patients in the rivaroxaban group compared to 351 of 18878 (1.85%) patients in the warfarin group (RR 0.76; 95% CI, 0.55, 1.06; heterogeneity I2 = 35%, P = 0.10), major bleeding in 247 of 4258 (5.8%) patients in the rivaroxaban group compared to 270 of 18879 (1.4%) patients in the warfarin group (HR 1.68; 95% CI, 0.59, 4.77; heterogeneity I2 = 97%) and intracranial hemorrhage in 24 out of 2583 (0.9%) patients in the rivaroxaban group compared to 35 of 2160 (1.6%) in warfarin group (HR 0.49; 95% CI, 0.16, 1.56; heterogeneity I2 = 70%).

**Conclusions:** Our results show that rivaroxaban is comparable to warfarin in prevention of stroke and systemic thromboembolism in patients with NVAF with VHD. Rivaroxaban is also comparable to warfarin in bleeding risks in these patients.

**REGISTRATION NUMBER:** CRD42021222490

## Introduction

The 2019 ACC/AHA Focused Updated on Atrial Fibrillation (AF) management defines valvular AF as AF in a patient with moderate-to-severe mitral stenosis or a mechanical heart valve and non-valvular AF (NVAF) as AF in the absence of moderate-to-severe mitral stenosis or a mechanical heart valve. NVAF with valvular heart disease (VHD) includes group of patients with NVAF and aortic stenosis or regurgitation, tricuspid valve stenosis or regurgitation, pulmonic stenosis or regurgitation, mitral regurgitation, mitral-valve prolapse, bioprosthetic valve and valve repair(1). Rivaroxaban and other novel oral anticoagulants (NOACs) are found to be safe and as efficacious as warfarin in patients with NVAF(2). However, we are lacking studies commenting in the role of individual NOAC in a NVAF with VHD. Many clinical trials that have studied the role of NOACs in NVAF have included patients with valvular heart disease (VHD) including bioprosthetic valve and valve repairs(3). With the completion of RIVER trail, published in November 2020, we attempted to perform a systematic review and meta-analysis to compare the safety and efficacy of individual NOAC, rivaroxaban against warfarin in prevention of thromboembolism in patients with NVAF with VHD. Our systematic review and meta-analysis include results from RIVER trail which was not included the previous reviews.

### Objectives

To compare the efficacy and safety of rivaroxaban against warfarin in patients with NVAF with VHD including bioprosthetic valve.

## Methods

### Eligibility Criteria

We included RCT, post-hoc of RCT and prospective and retrospective cohort studies in English language, studies reporting patients with valvular heart disease including bioprosthetic valvular disease. We searched both published and unpublished manuscripts and conference abstracts which fulfilled our eligibility criteria. Eligible studies compared rivaroxaban with warfarin in patients with NVAF and VHD and reported events of stroke, systemic embolism and major bleeding using appropriate definition. We excluded patients with valvular AF which includes moderate to severe mitral stenosis and mechanical heart valves, and hemodynamically unstable patients with valvular disease. We excluded patients with NVAF without VHD. Patients with rheumatic heart disease including those with moderate to severe mitral stenosis (MS) were excluded from the meta-analysis as being ineligible. Studies which did not report stroke and systemic embolism, major bleeding, and intracranial hemorrhage were deemed ineligible and not reported in our meta-analysis. We did not include case reports, case series, review articles and cross-sectional studies in our meta-analysis.

### Protocol and Registration

This systematic review was registered in the International prospective register of systematic reviews—PROSPERO (https://www.crd.york.ac.uk/PROSPERO/) with the registration reference CRD42021222490.

### Study Design

The Preferred Reporting Items for Systematic Reviews and Meta-Analyses (PRISMA) statement for reporting systematic reviews as recommended by the Cochrane Collaboration was followed in this systematic review(4).

### Information sources

A systematic literature review using MEDLINE, Embase, Cochrane CENTRAL, Scopus, and clinicaltrials.gov was done to search for articles published prior to April 2021.

### Search Strategy

We limited our search to studies conducted in humans, RCTs, post hoc of RCT and cohort studies. Search was restricted to English language. We searched the reference and citations of included articles and similar meta-analysis for additional articles. Search was performed using the terms “Rivaroxaban”, “Warfarin”, “Vitamin K antagonist”, “Oral Anticoagulants”, “Atrial Fibrillation”, “Valvular Heart Disease”, “Valve repair”, “Non-valvular Atrial Fibrillation”, and “Bioprosthetic Valve”. We search the above mesh terms for search. They were numbered #1 to #9. Then we combined them using “AND” and “OR” to get the final results.

### Data collection process

Search results were saved in EndNote version X9 (Developer: Clarivate analysis) files and transferred into Covidence software(5). We extracted the data manually through full text review.

### Selection and Data collection Process

Two reviewers independently performed the title and abstract screening and full text screening. Conflicts were resolved through consensus and if not resolved, third author resolved the conflict. Data collection was done by two authors (NB and RR).

### Data Items

All the studies were compatible to each outcome domain. All the studies compared rivaroxaban with warfarin in NVAF and VHD patients. The outcome of stroke and systemic embolism was most important as it was our efficacy outcome for comparing rivaroxaban against warfarin. We have defined the data items in Supplemental Table 1.

### Methodical quality assessment

We assessed the methodical rigor of the included studies using the modified Downs and Black checklist for RCTs and non-randomized studies(6). The checklist has 27 items with a total possible score of 28. Papers were rated excellent if they scored above 25, good if they scored between 20 and 25, fair if they scored between 15 and 19 and poor if they scored <15. Quality of studies and Risk of Bias of RCTs has been assessed in table 1. Each study was assessed by two independent investigators and discrepancies in scoring were resolved using consensus.

**Table 1:** Baseline characteristics of included studies. R =Rivaroxaban W= Warfarin AF= Atrial Fibrillation MS= Mitral Stenosis AS= Aortic Stenosis AR= Aortic Regurgitation PMB= Previous Major Bleeding Stroke/SE= Previous Stroke/ Systemic Embolism

| Study | Number of participants | Study design | Country | Study characteristics | Age |  |
| --- | --- | --- | --- | --- | --- | --- |
|  |  |  |  |  | R | W |
| Strange 2020(11) | 1735 | Observational, Prospective Cohort Study | Denmark | Danish Nationwide registry in patients with AF and Aortic stenosis/insufficiency, Mitral insufficiency, Bioprosthetic valves, Mitral and aortic valve repair | 80 | 77 |
| Briasoulis 2018(10) | 16158 | Observational Retrospective Cohort | USA | Medicare beneficiaries (age > 65 years) in patients with newly diagnosed AF and valvular disease included all valvular pathologies like MS, MR, TS, TR, AS, AR excluding Rheumatic MS, Bioprosthetic and Mechanical valve. | 77 | 80 |
| Breithardt 2014(8) | 14264 | Post hoc analysis of Randomized Double Blind Controlled Trial, both non-inferior and superior design | Multinational (45 nations) | Patient with AF and other valvular pathologies excluding Rheumatic and Mechanical Valve Stenosis or Prosthetic heart valve. | 73 | 73 |
| Guimarães 2020(9) | 1005 | Randomized Controlled Trial open label, non-inferiority trial design | Brazil | Patients with age $\geq 18$ years with AF and Bioprosthetic valve only were included excluding all other valvular lesions and mechanical valve. | 59.4 | 59.2 |

### Effect measure

#### Measure of Outcome

The efficacy outcome of interest were composites of stroke (ischemic, hemorrhagic and undetermined stroke) and systemic embolism.

The safety outcome were major bleeding and intracranial hemorrhage (ICH)(7).

#### Effect measure

Since, we pooled data from Cohort studies as well as RCTs, we used risk ratio as the effect measure for stroke and systemic embolism, major bleeding and ICH. 95% Confidence interval and p-value less than 0.05 was used to define the result as significant.

### Statistical analysis and synthesis method

Outcomes from the individual studies were aggregated with RevMan (version 5.4, Cochrane Collaboration, Oxford, United Kingdom) applying the Mantel-Haenszel test. Hazard ratio (HR) and 95% confidence intervals (CIs) were estimated using a fixed-effects model and random-effects method to account for the presence of variability among the studies. I^2^ statistic was used to assess heterogeneity. We did not perform meta-regression when heterogeneity was high. We just reported the heterogeneity.

To define heterogeneity, we used I^2^ index with values of ≤25%, 25% to 50%, and ≥50% were defined as low, moderate, and high degrees of heterogeneity. We used forest plot to graph the pooled results from analysis. We used random-effect model. Two-tailed p-values <.05 were considered to indicate statistical significance.

### Sensitivity analysis

Sensitivity analysis performed excluding retrospective and prospective cohort studies. However, as no significant change was observed we did not perform detail analysis. Reporting bias and certainty assessment: We did not have any missing results to perform the reporting bias. We did not use any tool for assessing the certainty.

### Publication Bias

We have only four studies included in the meta-analysis. We constructed forest plot for publication bias.

## RESULTS

### Study selection

We identified 122 articles from rom PubMed/MEDLINE and 3 articles from Embase and Web of Science (Figure 1). Duplicate studies were removed by the software. We identified 9 studies. Finally, 9 articles were fully read and 5 articles removed for not meeting eligibility. Final qualitative and quantitative analysis was done with 4 studies (Figure 1). [Insert figure 1]

**Figure 1:**
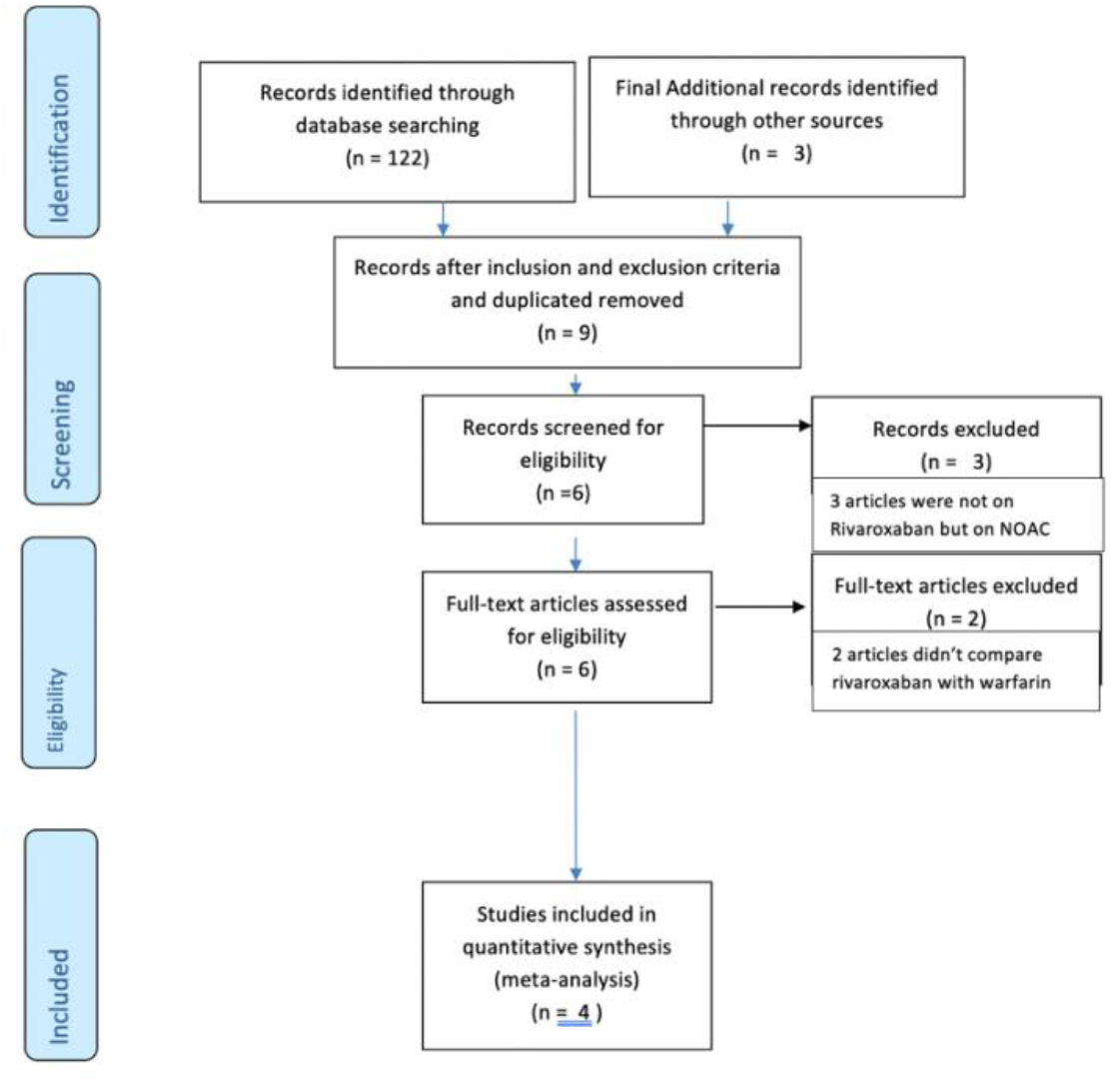
Prisma flow diagram of Included Studies

### Baseline Characteristics of Studies

The study by Breithardt et al. is a post hoc analysis of the ROCKET-AF trial(8). The study by Guimarães et al is part of the RIVER trail(9). The duration of follow up was 1.9 years and 1 year for ROCKET-AF and or RIVER trials, respectively. The study by Briasoulis et al. is a retrospective cohort analysis of claims data for adult Medicare beneficiaries age greater than 65 years(10). The study by Strange et al. is a nationwide cohort study of 2 years’ duration from January 1, 2014 until June 30, 2017. This study identified data from nationwide Danish registries with follow up until death, event, migration, switch, or discontinuation of treatment(11). The baseline characteristics of included studies were extracted and has been listed in table 1.

### CHADVaSc

(congestive heart failure, hypertension, age ≥ 75 years, diabetes mellitus, stroke or transient ischemic attack (TIA), vascular disease, age 65 to 74 years, sex category) score

### HASBLED

(Hypertension, Abnormal Renal/Liver Function, Stroke, Bleeding History or Predisposition, Labile INR, Elderly, Drugs/Alcohol Concomitantly) Score

### Risk of Bias and Quality of included studies

The results of the modified downs and black questionnaire tool are descripted in the table below. All included studies were rated either Good (2 Cohort studies) with score of 20/28 or Excellent (2 RCTs and post hoc of RCT) with score of 27/28 and 26/28 based on the modified Downs and Black criteria(6). Table 2.

**Table 2:**
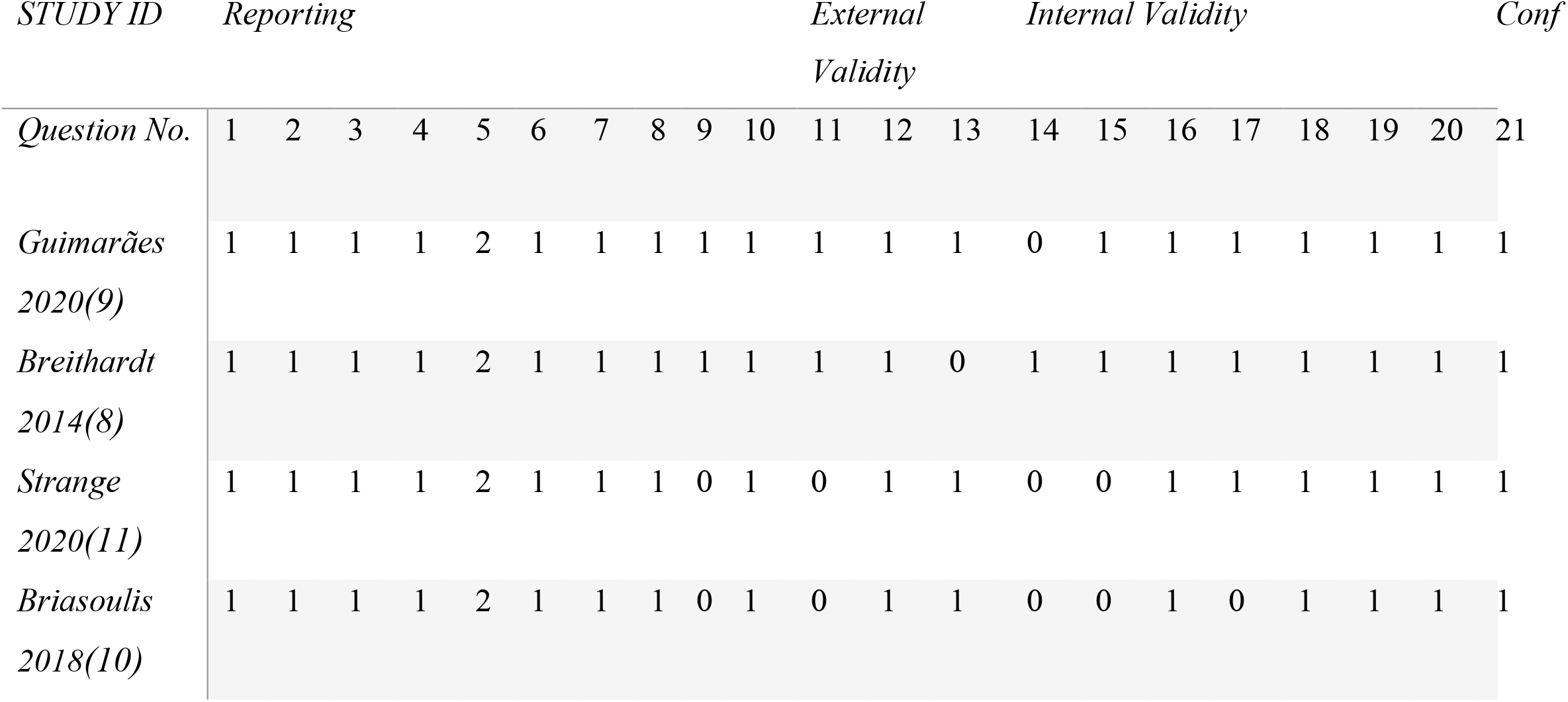
Quality of Included Studies P: Power Q: Quality of studies Tot: Total points of the study *E:Excellent ПG: Good

### Publication Bias

Due to only inclusion of four studies for the meta-analysis, we were unable to comment on publication bias in our study.

### Stroke and Systemic Embolism

Pooled data from four studies showed Stroke and Systemic embolism in 88 of 4258 (2.06%) patients in the rivaroxaban group and 351 of 18878 (1.85%) patients in the warfarin group. The pooled result showed that rivaroxaban is comparable to warfarin in prevention of Stroke and Systemic Embolism in patients with NVAF and VHD (RR 0.76; 95% CI, 0.55, 1.06; heterogeneity I^2^ = 35%, P = 0.10). (Figure 2)

**Figure 2:**
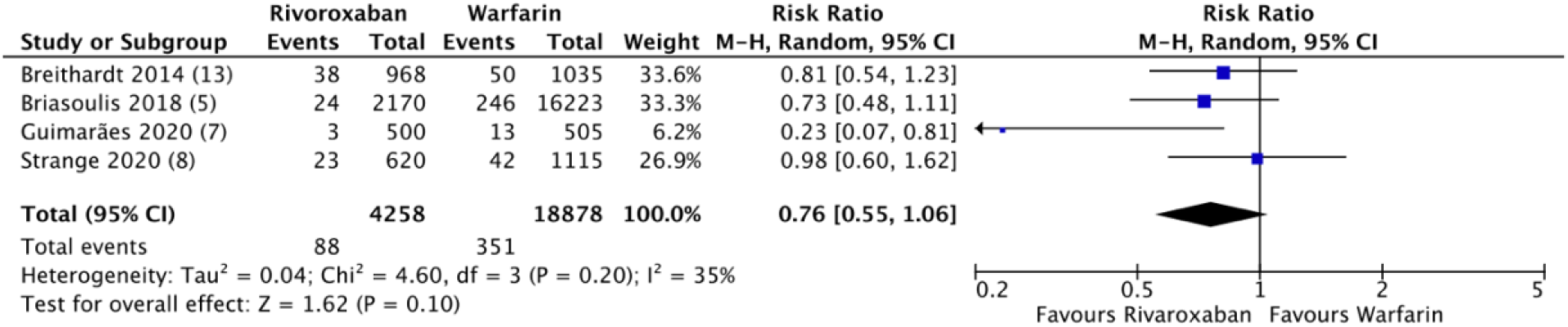
Forest plot showing comparison of stroke/systemic embolism between rivaroxaban and warfarin

### Major Bleeding

Pooled data from 4 studies showed Major bleeding in 247 of 4258 (5.8%) patients in the rivaroxaban group and 270 of 18879 (1.4%) patients in the warfarin group. Our results show that, there is no significant difference in major bleeding in the rivaroxaban group compared to warfarin. We report a high degree of heterogeneity in the outcome, among these studies (RR 1.68; 95% CI, 0.59, 4.77; heterogeneity I^2^ = 97%). (Figure 3)

**Figure 3:**
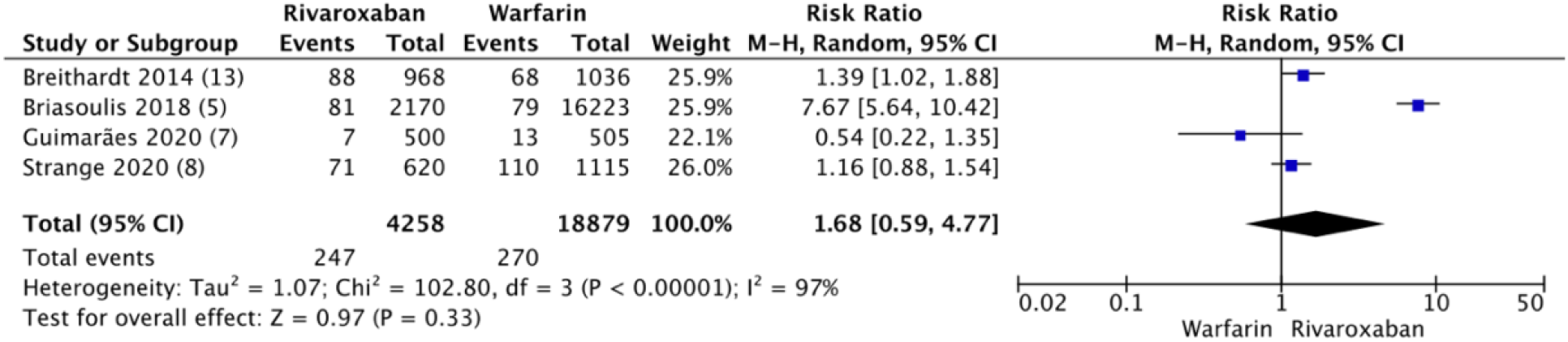
Forest plot showing comparison of Major Bleeding between rivaroxaban and warfarin

### Intracranial hemorrhage

Secondary safety outcome was ICH and the pooled data from 3 studies (as Study by Briasoulis et al(10) did not report ICH) showed the events of ICH in rivaroxaban were 24 out of 2583 (0.9%) compared to 35 of 2160 (1.6%) in warfarin group. Composing the data from 3 studies, the pooled result showed that, there is no significant difference in ICH in the rivaroxaban group compared to warfarin. We report a high degree of heterogeneity in the outcome, among these studies (RR 0.49; 95% CI, 0.16, 1.56; heterogeneity I^2^ = 70%). (Figure 4)

**Figure 4:**
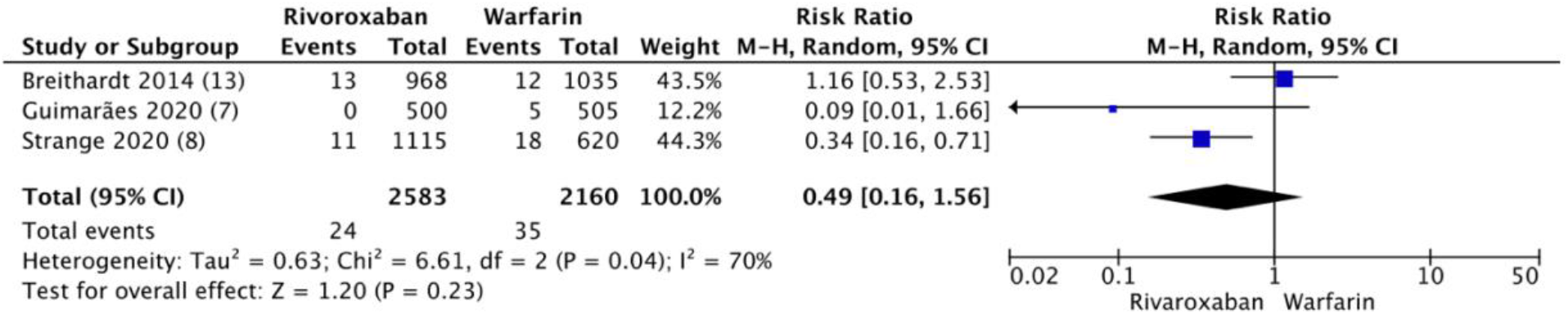
Forest plot showing comparison of Intracranial hemorrhage between rivaroxaban and warfarin

When analysis was limited to RCT or post hoc analysis of RCT only, results were similar (supplementary figures 1, 2, and 3 in Supplementary Material file).

## Discussion

### Stroke, Systemic Embolism, and ICH

Our study highlights that rivaroxaban is comparable to warfarin in prevention of stroke and systemic thromboembolism in patients with NVAF with VHD. Rivaroxaban is also comparable to warfarin in bleeding risks in these patients. The meta-analysis by Pan et al on NOACs versus warfarin showed reduced risk of stroke or systemic embolism (HR: 0.70; 95% CI, 0.60–0.82) and intracranial hemorrhage (HR: 0.47; 95% CI, 0.24–0.92) in NVAF patients with VHD. The study by Pan et al showed higher risk of major bleed with rivaroxaban (HR for major bleeding: 1.56 (95% CI, 1.20–2.04) and comparable risk of intracranial hemorrhage with 1.27 [95% CI, 0.77–2.10] (12). It is similar to our study in reporting non inferiority of rivaroxaban in preventing stroke and systemic embolism and comparable risk of ICH. However, the Pan et al differs from our study in reporting higher risk of major bleeding in rivaroxaban group whereas our study showed comparable effect. The inclusion of cohort study by Briasoulis et al significantly affected our results as the RR of the study is 7.67 [95% CI, 5.64, 10.42]. This study is subject to high risk of bias because of its cohort study design.

Caldeira et al found similar findings of decreased risk of stroke and SE while comparing NOACs with warfarin in patients with NVAF and VHD with HR 0.73, (95% CI: 0.60–0.90)(13). Caldeira et al reported NOACs did not show significantly different risk of major bleeding in patients with NVAF and VHD compared with warfain (HR 0.91, 95% CI 0.68–1.23) but I^2^ is 71% showing high statistical heterogeneity. This finding is somewhat similar to our study. However, the study showed significant risk reduction of ICH in VHD patients (HR 0.45, 95% CI 0.24–0.87) which contrast with our study but the outcome is highly heterogenous with I^2^ of 69%(13).

To date, our study is the only study that has examined the safety and efficacy profile of the individual NOAC, rivaroxaban. While aggregate results of all NOACs is helpful, clinicians need individual NOAC’s data for prescribing.

The 2019 AHA/ACC/HRS (American Heart Association/American College of Cardiology/Heart and Rhythm Society) guidelines recommend the use of warfarin and NOACs for the treatment of NVAF, while warfarin alone is recommended for patients with AF and mechanical heart valves. It defines “NOAC-eligible” patients as those with the absence of moderate to severe mitral stenosis or a mechanical heart valve(1).

In a study by Mekaj et al, NOACs have lower incidence of major bleeding, convenience of use, minor drug and food interactions, a wide therapeutic window, and no need for laboratory monitoring which further favors NOACs including rivaroxaban(14).

### Limitations

Two of our selected studies were non-RCTs and one was a post hoc of RCT and two were observational cohort studies. Therefore, the problem of selection bias and confounding is a major limitation. There is wide confidence interval which suggest that the study many not have enough power even when if there is a real difference (Type II error). There is high statistical heterogeneity in the outcomes of major bleeding and ICH in our study which we were unable to address in our study. In the warfarin arm, time to therapeutic range was unknown due to missing long-term INR data. We were unable to ascertain the impact of comorbidities on both safety and efficacy outcomes. We were unable to include all-cause mortality due to lack of data in all the four studies. The quality of studies assessed by the Downs and Black tool shows that all the included studies were not excellent in quality which could also be a limitation our study(6). Due to lack of RCTs directly comparing Rivaroxaban with Warfarin, we had to include observational studies comparing rivaroxaban with warfarin.

## Conclusions

Our results show that rivaroxaban is comparable to warfarin in prevention of stroke and systemic thromboembolism in patients with NVAF with VHD. Rivaroxaban is also comparable to warfarin in bleeding risks.

## Future Perspectives

We are lacking meta-analysis on individual NOAC as all NOAC are not the same in safety and efficacy. Most of the published meta-analysis included all the NOAC in the same study which may be prone to bias. We are lacking enough RCTs to compare individual NOACs.

## Data Availability

The data are provided from all the individuals studies referenced in the journal.

## SUPPLEMENTAL FILES

**Supplemental Table 1:** Definition of data outcomes and time frame for included studies.

| Study | Strange 2020<sup>6</sup> | Briasoulis 2018<sup>7</sup> | Guimarães 2020<sup>8</sup> |
| --- | --- | --- | --- |
| Defining efficacy outcome of stroke and systemic embolism | Hospitalizations with ischemic stroke, unspecified stroke, systemic embolism, or TIA captured with ICD10 code: I63-64, I74, G458-459 | Hospitalizations with ischemic stroke, unspecified stroke, systemic embolism, or TIA captured with ICD 9 CMcode | Non-traumatic, focal and sudden resulting from Cerebrovascular ca within 24 hours, no readily identified tumor or seizure, or which resolves accompanied by clear evidence of imaging. Systemic embolism as associated with clinical and radiologic arterial occlusion with or without mechanism e.g. trauma, at instrumentation. |
| Defining safety outcome major bleed | Defined from ICD10 codesD500-S368D | Defined from ICD9CM code | Defined same as in ROCKET_AF t Clinically overt bleeding associated outcome, involving a critical site, or bleeding associated with a fall in hemoglobin concentration of $\geq 2$ g/dL, or leading to $\geq 2$ units of packed red blood cells or TIMI criteria or BARC criteria. |
| Defining safety outcome<br>Intracranial hemorrhage | Defined from ICD 10 code I60-62, SO64-66 | Defined from ICD9 CM code | Same as ROCKET-AF or TIMI or |
| Time frame | 42 months observational data from danish national registry | 24 months retrospective data | 12 months |
| Defining Atrial Fibrillation | Defined from ICD 10 code: I48 | Defined from ICD 9 CM code 427.31 for new onset AF | Same as ROCKET AF trial |

## SUPPLEMENTARY Material

Outcomes in Randomized Controlled Trials only:

### Stroke and Systemic Embolism in RCTs

Pooled data from one RCT and one post hoc of RCT showed Stroke and Systemic embolism in 27 of 2670 (1.01%) patients in the rivaroxaban group and 259 of 16728 (1.54%) patients in the warfarin group. The pooled result showed that there was non-significant reduction in incidence of Stroke and Systemic Embolism in patients taking rivaroxaban versus warfarin (HR 0.48; 95% CI, 0.16, 1.42; heterogeneity I^2^ = 65%). Supplementary Figure 1.

**Supplementary figure 1:**
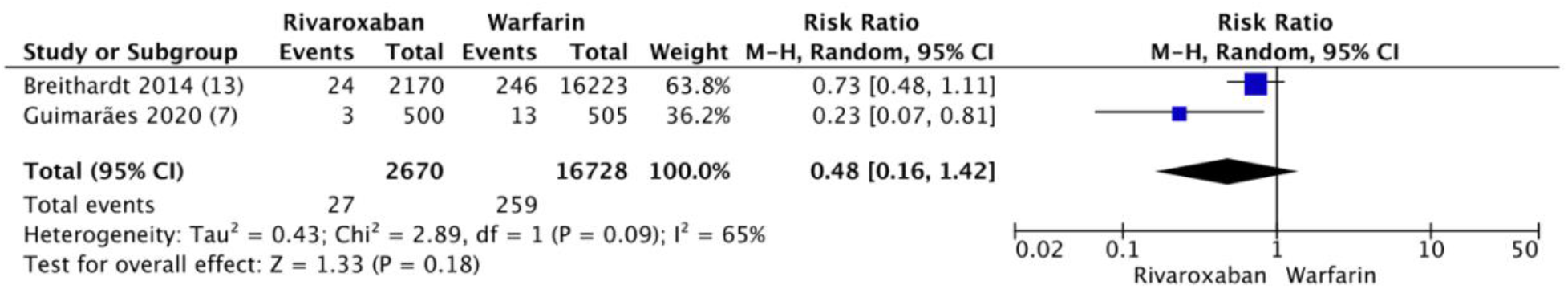
Forest plot showing comparison of stroke/systemic embolism between rivaroxaban and warfarin in RCTs

### Major Bleeding

Pooled data from one RCTs and one post hoc of RCT showed Major bleeding in 95 of 1468 (6.4%) patients in the rivaroxaban group and 81 of 1541 (5.2%) patients in the warfarin group. The pooled result showed that although the incidence of major bleeding was higher in the rivaroxaban group compared to warfarin, the difference was not statistically significant. There was, however, a high degree of heterogeneity among these studies (HR 0.96; 95% CI, 0.39, 2.35; heterogeneity I^2^ = 73%). Supplementary Figure 2.

**Supplementary figure 2:**
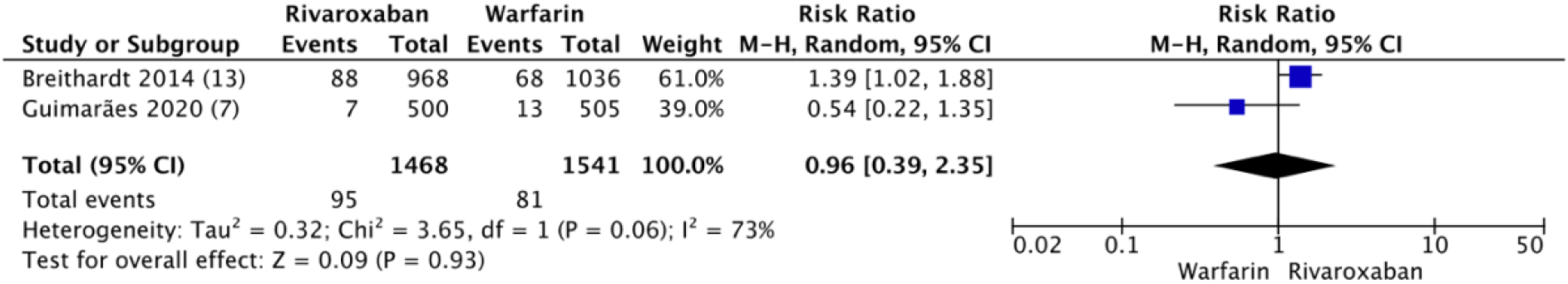
Forest plot showing comparison of major bleeding between rivaroxaban and warfarin in RCTs

Intracranial hemorrhage:

Pooled data from one RCTs and one post hoc of RCT showed the events of ICH in rivaroxaban 13 out of 1468 (0.88%) compared to 17 of 1540 (1.1%) in warfarin group. Composing the data from 2 RCTs, the pooled result showed that there was non-significant reduction in incidence of intracranial hemorrhage in the patients taking rivaroxaban versus warfarin (HR 0.47; 95% CI, 0.04, 5.56; heterogeneity I^2^ = 66%). Supplementary Figure 3.

**Supplementary figure 3:**
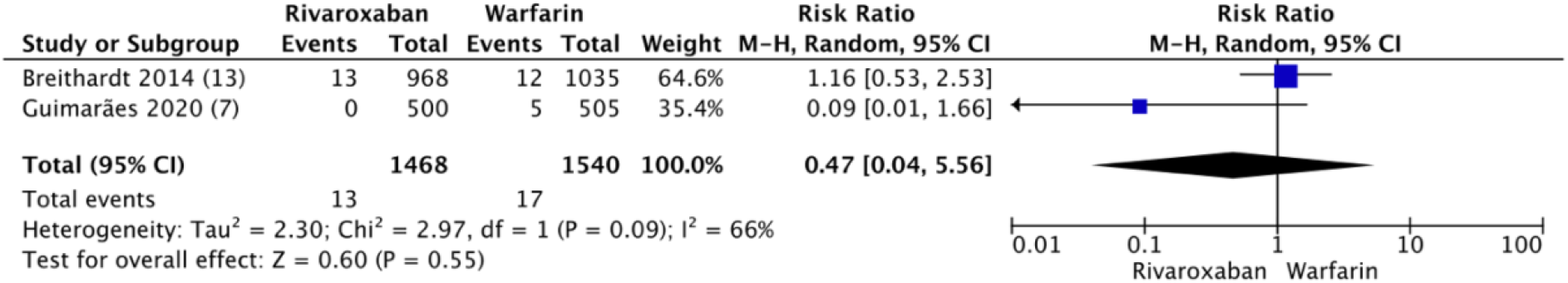
Forest plot showing comparison of intracranial hemorrhage between rivaroxaban and warfarin in RCT

## References

1. January CT, Wann LS, Calkins H, Chen LY, Cigarroa JE, Cleveland JC, Jr., et al. 2019 AHA/ACC/HRS Focused Update of the 2014 AHA/ACC/HRS Guideline for the Management of Patients With Atrial Fibrillation: A Report of the American College of Cardiology/American Heart Association Task Force on Clinical Practice Guidelines and the Heart Rhythm Society. J Am Coll Cardiol. 2019;74(1):104–32.

2. Hampton ML, Tellor KB, Armbruster AL, Theodos G, Schwarze MW. Evaluation of the Safety and Effectiveness of Direct-Acting Oral Anticoagulants in Patients with Atrial Fibrillation and Coexisting Valvular Heart Disease. Am J Cardiovasc Drugs. 2020;20(6):611–7.

3. Ruff CT, Giugliano RP, Braunwald E, Hoffman EB, Deenadayalu N, Ezekowitz MD, et al. Comparison of the efficacy and safety of new oral anticoagulants with warfarin in patients with atrial fibrillation: a meta-analysis of randomised trials. Lancet. 2014;383(9921):955–62.

4. Moher D, Liberati A, Tetzlaff J, Altman DG. Preferred reporting items for systematic reviews and meta-analyses: the PRISMA statement. PLoS Med. 2009;6(7):e1000097.

5. COVIDENCE [Internet]. 2021. Available from: https://app.covidence.org/reviews/active.

6. Downs SH, Black N. The feasibility of creating a checklist for the assessment of the methodological quality both of randomised and non-randomised studies of health care interventions. J Epidemiol Community Health. 1998;52(6):377–84.

7. Schulman S, Kearon C. Definition of major bleeding in clinical investigations of antihemostatic medicinal products in non-surgical patients. J Thromb Haemost. 2005;3(4):692–4.

8. Breithardt G, Baumgartner H, Berkowitz SD, Hellkamp AS, Piccini JP, Stevens SR, et al. Clinical characteristics and outcomes with rivaroxaban vs. warfarin in patients with non-valvular atrial fibrillation but underlying native mitral and aortic valve disease participating in the ROCKET AF trial. Eur Heart J. 2014;35(47):3377–85.

9. Guimarães HP, Lopes RD, de Barros ESPGM, Liporace IL, Sampaio RO, Tarasoutchi F, et al. Rivaroxaban in Patients with Atrial Fibrillation and a Bioprosthetic Mitral Valve. N Engl J Med. 2020;383(22):2117–26.

10. Briasoulis A, Inampudi C, Akintoye E, Alvarez P, Panaich S, Vaughan-Sarrazin M. Safety and Efficacy of Novel Oral Anticoagulants Versus Warfarin in Medicare Beneficiaries With Atrial Fibrillation and Valvular Heart Disease. J Am Heart Assoc. 2018;7(8).

11. Strange JE, Sindet-Pedersen C, Staerk L, Grove EL, Gerds TA, Torp-Pedersen C, et al. All-cause mortality, stroke, and bleeding in patients with atrial fibrillation and valvular heart disease. Eur Heart J Cardiovasc Pharmacother. 2020.

12. Pan KL, Singer DE, Ovbiagele B, Wu YL, Ahmed MA, Lee M. Effects of Non-Vitamin K Antagonist Oral Anticoagulants Versus Warfarin in Patients With Atrial Fibrillation and Valvular Heart Disease: A Systematic Review and Meta-Analysis. J Am Heart Assoc. 2017;6(7).

13. Caldeira D, David C, Costa J, Ferreira JJ, Pinto FJ. Non-vitamin K antagonist oral anticoagulants in patients with atrial fibrillation and valvular heart disease: systematic review and meta-analysis. Eur Heart J Cardiovasc Pharmacother. 2018;4(2):111–8.

14. Mekaj YH, Mekaj AY, Duci SB, Miftari EI. New oral anticoagulants: their advantages and disadvantages compared with vitamin K antagonists in the prevention and treatment of patients with thromboembolic events. Ther Clin Risk Manag. 2015;11:967–77.

